# MY008211A in complement inhibitor–naive patients with paroxysmal nocturnal hemoglobinuria and signs of hemolysis

**DOI:** 10.64898/2026.02.05.26345159

**Authors:** Lei Ye, Miao Chen, Hongyan Tong, Bing Han, Li Zhang, Hong Chang, Xin Li, Zengmei Sheng, Chen Yang, Gaixiang Xu, Nina Guo, Yongkai Chen, Rui Xia, Chunlan Tang, Li Liu, Xiaodan Guo, Yihan Zhang, Xiaoni Li, Rito Ki, Wilson Chunlin Wang, Geoffrey Ross, Carlos de Castro, Chuanrui Xu, Fengkui Zhang

## Abstract

**Key points:**

- We report findings from a phase 2 study of MY008211A among Chinese men and women aged ≥18 years with paroxysmal nocturnal hemoglobinuria
- Increases in hemoglobin of ≥20 g/L were maintained for up to 44 weeks of treatment with MY008211A in all 34 patients^iv^

**Explanation of novelty:** Paroxysmal nocturnal hemoglobinuria is characterized by red blood cell (RBC) destruction and a prothrombotic state.^v^ Treatments exist such as complement 5 inhibitors but these carry the risk for iatrogenic extravascular hemolysis and anemia.^vi^ As reported here, the novel, oral complement factor B inhibitor MY008211A yielded increases in hemoglobin and RBC levels, while adverse events over 44 weeks were largely mild to moderate in severity, and infections generally consisted of respiratory infections.^vii^

Paroxysmal nocturnal hemoglobinuria (PNH) is a life-threatening disease characterized by red blood cell (RBC) destruction, blood clots, and impaired bone marrow function.^viii^ We evaluated the efficacy and safety of 3 dosages of MY008211A, a novel complement factor B inhibitor,^ix^ for treating PNH.^x^ This was a multicenter, open-label, phase 2, dose-finding study of MY008211A among Chinese men and women with complement inhibitor–naive PNH and signs of active hemolysis.^xi^ Patients with hemoglobin <100 g/L were assigned to oral MY008211A 400 mg twice daily (BID), 600 mg BID, or 800 mg once daily (QD) for 12 weeks and could then continue treatment with 400 mg BID during a 32-week extension.^xii^ The primary endpoint was the proportion of patients achieving an increase in hemoglobin concentration of ≥20 g/L vs baseline on day (D)84, without RBC transfusions after 4 weeks of dosing.^xiii^ Safety assessments included adverse events (AEs).^xiv^ Fifteen, 9, and 10 patients were assigned to MY008211A 400 mg BID, 600 mg BID, and 800 mg QD, respectively.^xv^ All patients completed the study and its 32-week extension.^xvi^ On D84, all 34 patients achieved increases in hemoglobin concentration of ≥20 g/L from baseline;^xvii^ all patients maintained this increase at D308.^xviii^ Through D308, grade ≥3 AEs occurred in 5 (33%), 5 (56%), and 4 (40%) patients in the 400-, 600-, and 800-mg groups, respectively.^xix^ There were no deaths.^xx^ In this multicenter, open-label study of 3 dosages of MY008211A for PNH, all patients achieved and maintained increases in hemoglobin of ≥20 g/L from baseline without RBC transfusions.

## Introduction

Paroxysmal nocturnal hemoglobinuria (PNH) is a life-threatening disease characterized by red blood cell (RBC) destruction, blood clots, and impaired bone marrow function.^1xxi^ PNH causes vasoconstriction and platelet activation and leads to renal insufficiency and pulmonary hypertension, creating a dangerously prothrombotic environment.^2xxii^ Its estimated prevalence is 3.81 per 100,000 persons.^3xxiii^

Current options for management of PNH include the complement 5 (C5) inhibitors eculizumab, ravulizumab, crovalimab, and pozelimab; the complement 3 inhibitor pegcetacoplan; the complement factor D inhibitor danicopan, which is given in combination with a C5 inhibitor^4xxiv^; and the complement factor B inhibitor iptacopan.^4xxv^ These noncurative therapies have noteworthy drawbacks; C5 inhibition presents risks for iatrogenic extravascular hemolysis and persistent anemia, contributing to a continued need for transfusion,^4–6xxvi^ and the required intravenous administration of several agents is inconvenient and often poorly tolerated.^5xxvii^ Additionally, there are increased risks for both bacterial and viral infections associated with complement inhibition^7xxviii^ and breakthrough hemolysis from a complement-activating event.^6xxix^ The only potentially curative therapeutic modality for PNH is allogeneic hematopoietic stem cell transplant (allo-HSCT).^5xxx^ However, there are significant morbidity and mortality risks associated with this mode of therapy; one recent study reported 1-year rates of mortality greater than 10% and of chronic graft-vs-host disease of 23.5% or more.^8xxxi^ Thus, allo-HSCT is only a viable option in cases of complicated or life-threatening PNH disease, such as in high-risk myelodysplastic syndrome, patients with complications of PNH that are unresponsive to eculizumab, or patients with severe aplastic anemia who have an available matched donor.^9xxxii^ A broader range of therapeutic options are needed for patients for whom the benefits of transplant do not outweigh the risks.

## Methods

This study evaluated the efficacy and safety of 3 dosages of MY008211A, a complement factor B inhibitor, in the treatment of patients with PNH.

### Ethics

The study protocol was approved by the institutional review boards or independent ethics committees at each participating site, including the Institute of Hematology and Blood Diseases Hospital, Chinese Academy of Medical Sciences; Peking Union Medical College Hospital, Chinese Academy of Medical Sciences; The First Affiliated Hospital, Zhejiang University School of Medicine; West China Hospital, Sichuan University; and Changsha Third Hospital. The study was conducted in accordance with the principles of the Declaration of Helsinki and the International Council for Harmonisation Good Clinical Practice guidelines. All participants provided written informed consent prior to study participation.

Statistical analyses were performed by statisticians designated by the sponsor’s selected vendor. All authors had access to the primary clinical trial data and take responsibility for the integrity of the data and the accuracy of the data analysis.^xxxiii^ The trial was registered with clinicaltrials.gov (NCT06050226).^xxxiv^

### Study design

This was an open-label, phase 2, dose-finding study of MY008211A conducted at 5 centers in China.^xxxv^ Men and women aged ≥18 years with PNH and signs of active hemolysis were eligible (**Figure 1**).^xxxvi^ Diagnosis of PNH was based on China’s Rare Disease Diagnostic and Treatment Guidelines (2019 edition),^10xxxvii^ and patients were required to have PNH erythrocyte or granulocyte clone levels >10% (based on flow cytometry) within the 6 months prior to or during screening.^xxxviii^ Over the 8-week screening period, hemoglobin concentration was required to be <100 g/L at the first screening and treated with RBC transfusion for anemia due to PNH during screening, or recorded at a mean concentration <100 g/L in two assays separated by 2 to 8 weeks. At least two tests (2–8 weeks apart) during the screening period must have shown serum lactate dehydrogenase (LDH) values >1.5 × the upper limit of normal (ULN). Those who qualified for the study received meningococcal and *Streptococcus pneumoniae* vaccinations if they had not been vaccinated within 3 and 5 years, respectively.^xxxix^

**Figure 1.**
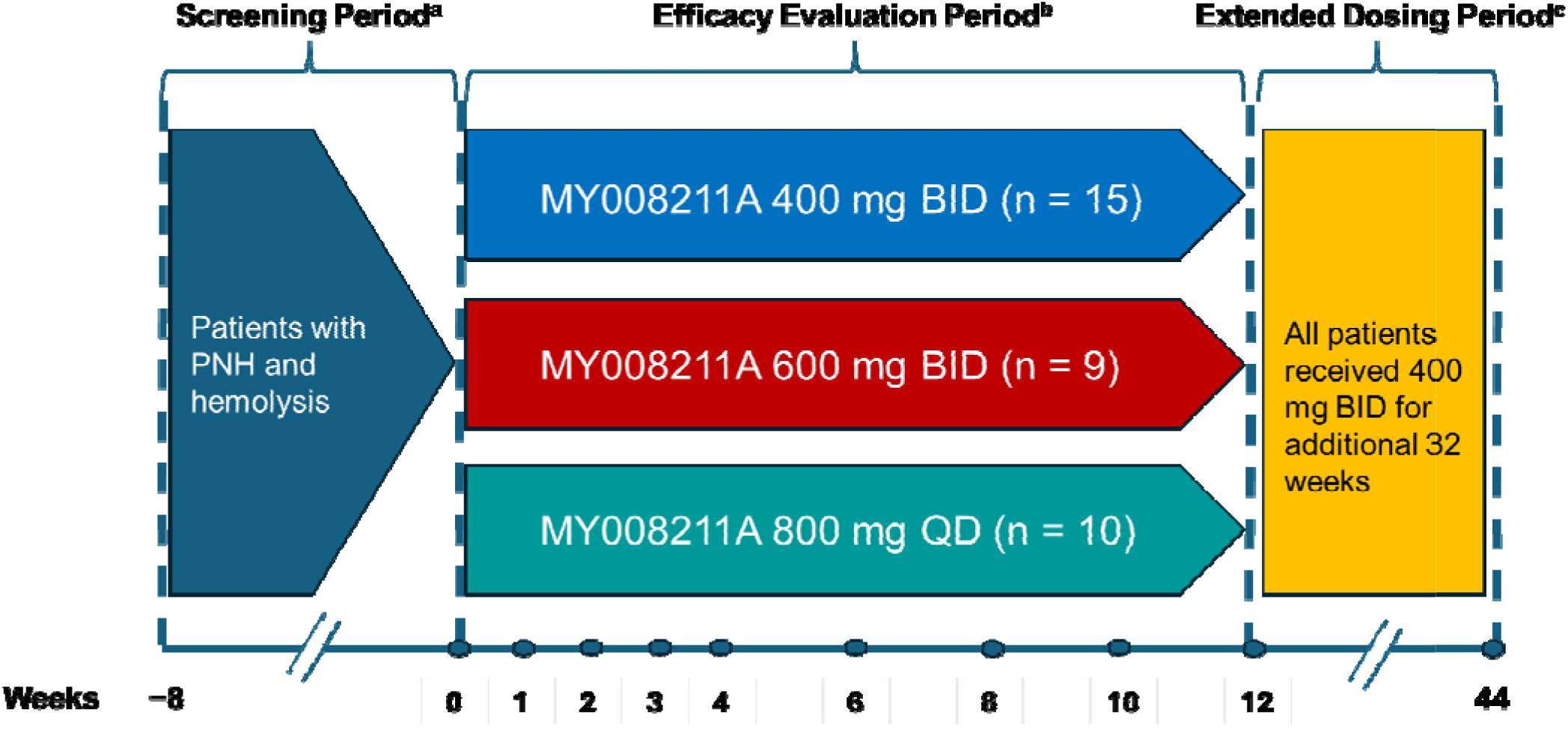
Study design^ci^. ^a^All candidates entering the screening phase were initially screened after completion and signing of informed consent at visit 1, and those patients who qualified for the initial screening (at the investigators’ discretion) were vaccinated. Screening and confirmation of entry and exclusion criteria were performed again at visit 2, and patients with PNH and active hemolysis who met the eligibility criteria were entered into the efficacy observation period. ^b^All enrolled patients were required to return to the study center at days 1, 7, 14, 21, 28, 42, 56, 70, and 84 after fasting for at least 6 hours to complete the relevant examinations. ^c^At the end of the efficacy observation period, if the investigators determined that the benefits of continuing to take the study drug outweighed the risks, the patients could enter the extended-dosing period. Patients entering the extended-dosing period continued or switched to the 400-mg BID regimen.^cii^ BID, twice daily; PNH, paroxysmal nocturnal hemoglobinuria; QD, once daily.

During the treatment phase, the first 10 patients enrolled were all assigned to receive MY008211A 400 mg twice daily (BID), the next 15 patients enrolled were randomized 1:2 to receive MY008211A 400 mg BID or 600 mg BID, and the last 10 patients enrolled were assigned to receive MY008211A 800 mg once daily (QD).^xl^ All patients took the study drug orally in a fasted or fed state at the same time every day whenever possible, morning and evening for patients assigned to 400 or 600 mg BID and once daily for those assigned to 800 mg QD, for a total of 12 weeks. Patients who entered the extended-dosing phase were switched to 400 mg BID for continued dosing over 32 weeks following a dose-response analysis.^xli^

The primary efficacy endpoint was the proportion of patients with an increase in hemoglobin concentration of ≥20 g/L on day 84 compared with baseline without RBC transfusions after 4 weeks of dosing.^xlii^ Secondary endpoints were as follows:^xliii^ the proportion of patients with an increase in hemoglobin concentration of ≥20 g/L from baseline without RBC transfusions (days 14, 21, 28, 42, 56, and 70); the proportion of patients with hemoglobin concentration ≥120 g/L without RBC transfusions, change from baseline in hemoglobin concentration in patients without RBC transfusions, and the proportion of patients without RBC transfusions (all measured at days 14, 21, 28, 42, 56, 70, and 84); change from baseline in serum LDH levels, the proportion of patients with hemolytic control (serum LDH <1.5 × ULN), change from baseline in reticulocyte counts in patients without RBC transfusions, change from baseline in serum indirect bilirubin levels, and change from baseline in Functional Assessment of Chronic Illness Therapy–Fatigue (FACIT-F) scores (all measured at days 7, 14, 21, 28, 42, 56, 70, and 84); change in average weekly RBC transfusions during the efficacy observation period compared with the pre-administration period; and change from baseline in PNH RBC clone levels in patients without RBC transfusions (measured on day 84).

Efficacy analyses were also conducted post hoc using data collected over the 32-week extension on days 112, 140, 168, 196, 224, 252, and 308, as follows:^xliv^ hemoglobin concentration increase ≥20 g/L without RBC transfusions, hemoglobin concentration ≥120 g/L without RBC transfusions, hemoglobin concentration and change from baseline, LDH concentration and change from baseline, the proportion of patients maintaining hemolytic control (defined as serum LDH <1.5 × ULN),^xlv^ reticulocyte count and change from baseline, RBC count and change from baseline, serum indirect bilirubin level and change from baseline, and the proportion of patients without RBC transfusion.^xlvi^

### Statistical methods^xlvii^

This study used a historical control design; we compared outcomes for the primary endpoint against the target value of the historical control to determine whether there was a significant difference between the two. Consulting hematologists had advised that the rate of hemoglobin increase of ≥20 g/L from baseline among patients with PNH not treated with complement inhibitors may be expected to be approximately 15%. The therapeutic efficacy of MY008211A tablets was expected to lead to such increases in more than 15% of patients. Therefore, we used a 1-sample rate superiority hypothesis test (H0: *P* ≤0.15, H1: *P* >0.15) using α = 0.025.

For the primary endpoint, 95% confidence intervals and *P*-values were calculated. Based on the suggested efficacy effect, using the sample size estimation method of a single-group target-value test with the assumption that at least 45% of patients would achieve the above-defined increase in hemoglobin concentration at day 84^xlviii^ and setting α = 0.025 and β = 0.2, the sample size of the highest-dosage group was calculated to be approximately 8 patients. Anticipating a 15% dropout rate, we determined that approximately 10 patients with PNH could be included in each group in the primary endpoint analysis. To more fully describe drug safety, we sought to enroll approximately 15 patients into the 400-mg BID group and approximately 10 each into the 600-mg BID and 800-mg QD groups, for a total sample size of approximately 35 patients.^xlix^ Analysis of secondary endpoints was conducted in a similar manner to that used for the primary endpoint. Safety events were summarized descriptively.^l^ The analyses for the extended-dosing period were considered exploratory. Results for these outcomes are presented as point estimates with 95% confidence intervals.^li^

## Results

### Study population

Fifteen patients were assigned to MY008211A 400 mg BID, nine to 600 mg BID, and ten to 800 mg QD.^lii^ Patient characteristics were similar across groups (**Table 1**).^liii^ Of the 34 patients, 15 were male, and 19 were female; the mean age was 39.5 years, and all patients were of Han ethnicity, except for 1 in the 800-mg QD group who was of Tujia Chinese ethnicity.^liv^ Mean (standard deviation [SD]) body mass index was 23.6 (3.86) kg/m^2^, and a history of aplastic anemia was present in 5 (33%), 5 (56%), and 4 (40%) patients in the 400-mg BID, 600-mg BID, and 800-mg QD groups, respectively.^lv^ All 34 patients completed the trial and extension (**Supplemental Figure 1**).^lvi^ Compliance with dosing was ≥98% in all dosage groups.^lvii^

**Table 1.**
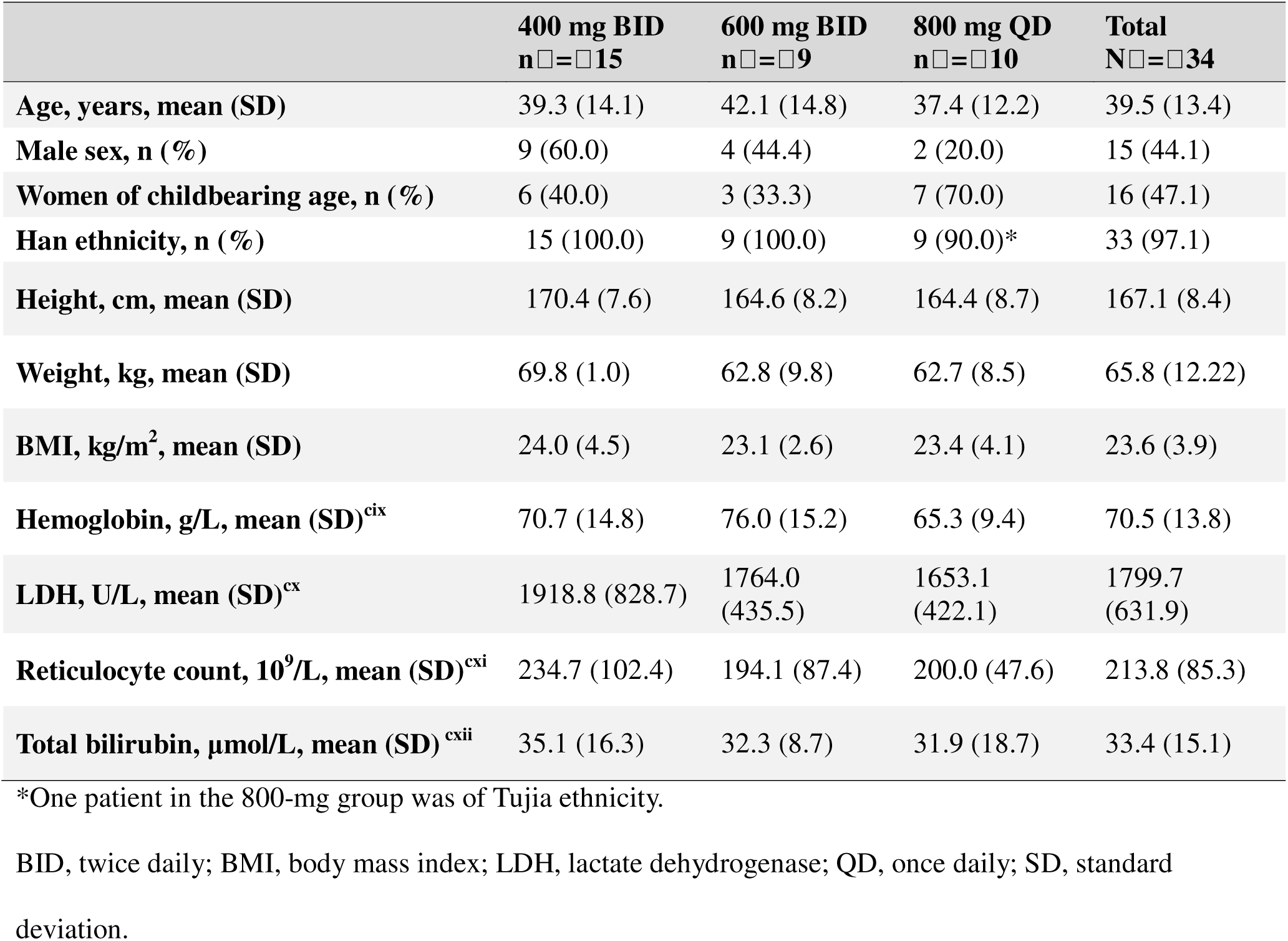
Baseline characteristics^cviii^.

### Efficacy

In the assessment of efficacy over the predefined 12-week efficacy evaluation period, all 34 patients treated with MY008211A had achieved the primary endpoint of an increase in hemoglobin concentration of ≥20 g/L from baseline at week 12 (**Figure 2a**).^lviii^ In the 400-mg BID group, all patients achieved this by week 8; in the 600-mg BID group, all patients achieved this by week 10; in both groups, 100% achievement was maintained at week 12. All 3 dosage groups showed significant differences vs the historical control (*P*[<0.0001).^lix^

**Figure 2.**
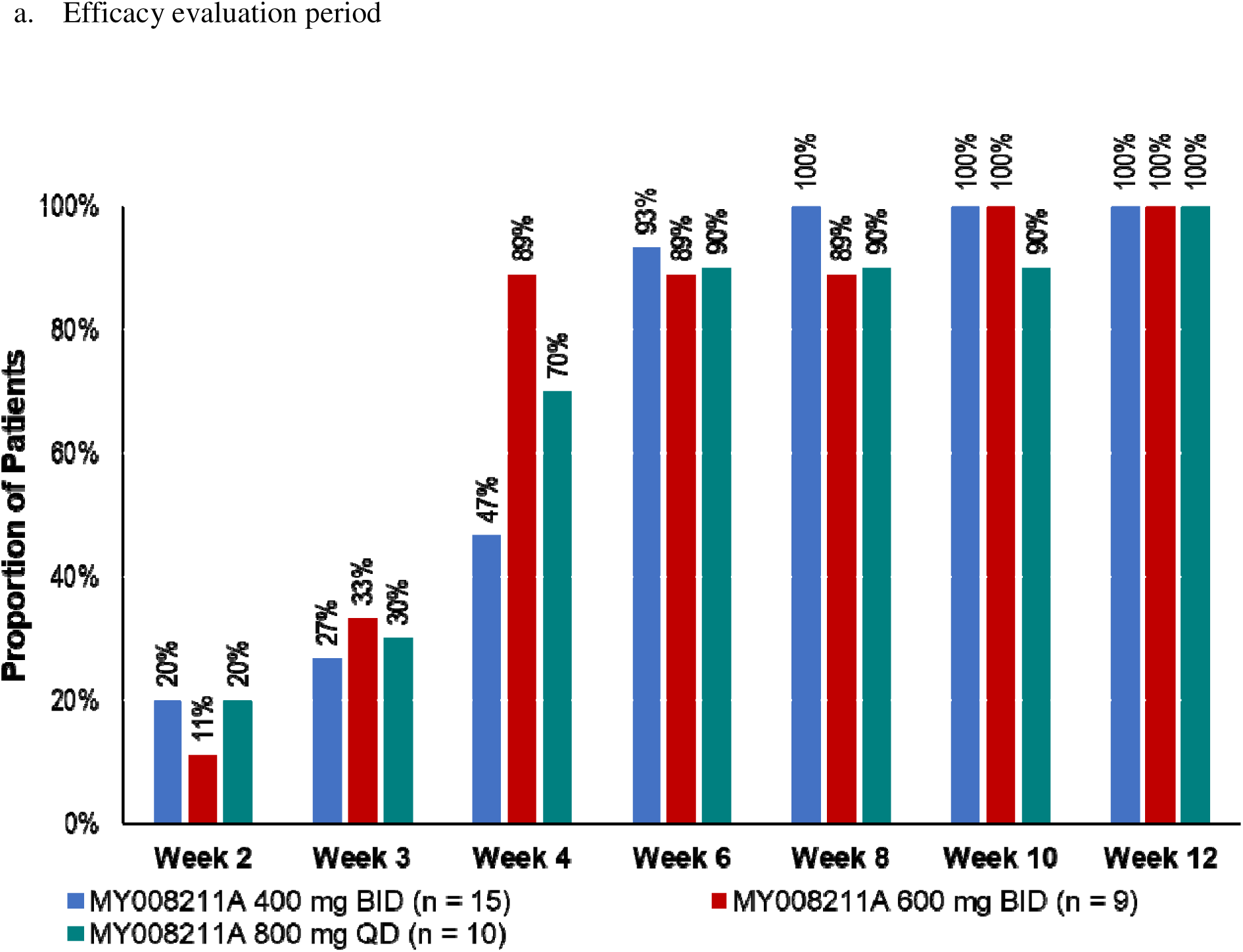
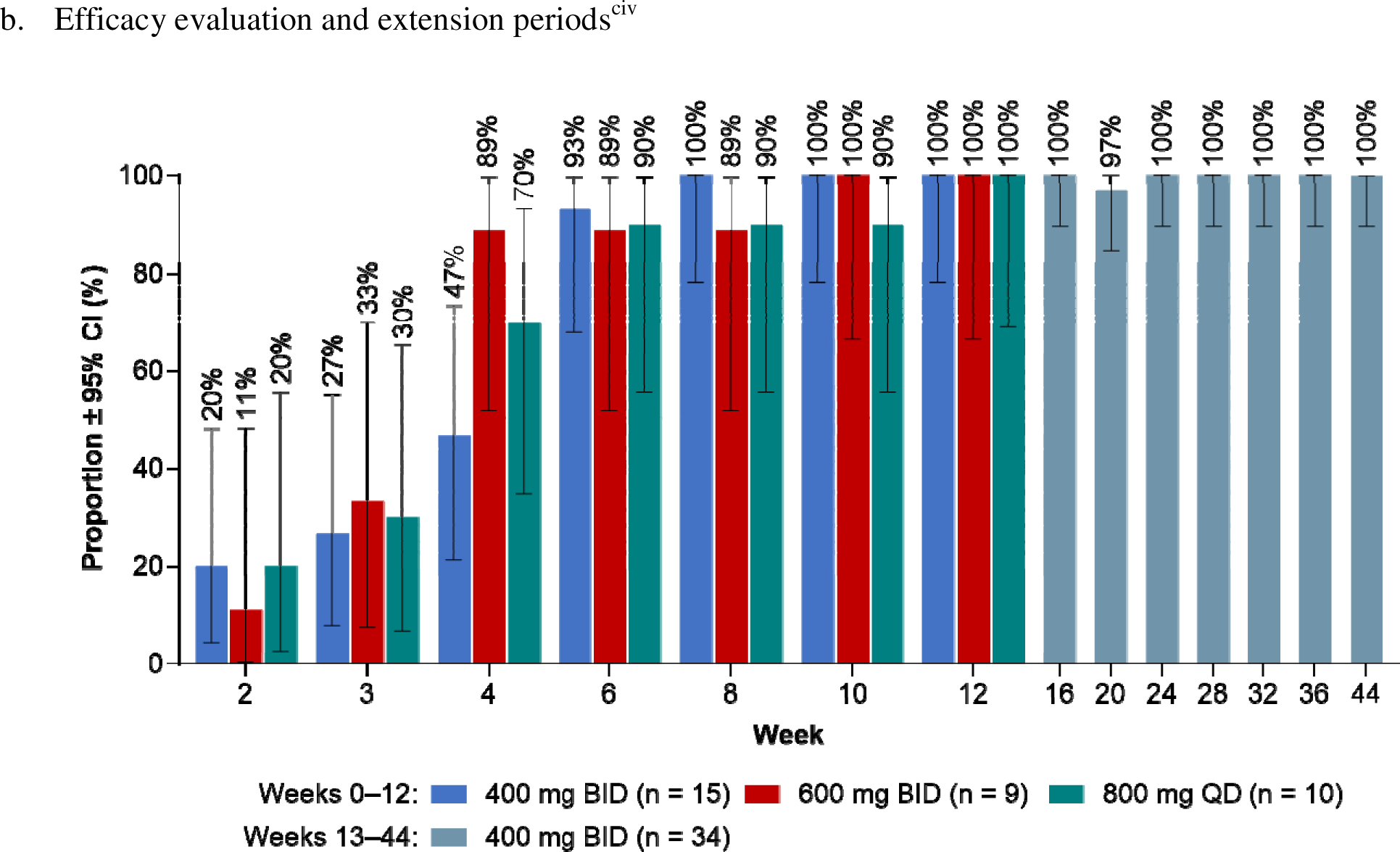
Proportions of patients with hemoglobin increases of ≥20 g/L from baseline with no red blood cell transfusion^ciii^. BID, twice daily; CI, confidence interval; QD, once daily.

The results of the secondary efficacy endpoints over the efficacy evaluation period were similar to those of the primary endpoint. Six of 15 patients (40%) treated with 400 mg BID, 4 of 9 (44%) treated with 600 mg BID, and 2 of 10 (20%) treated with 800 mg QD achieved a hemoglobin concentration ≥120 g/L (**Figure 3a**).^lx^ All treatment groups achieved significant increases in hemoglobin concentration vs baseline at week 12, with mean (SD) increases of 44.7 (12.98), 43.6 (13.21), and 42.4 (14.40) g/L in the 400-mg BID, 600-mg BID, and 800-mg QD groups (*P* <0.0001, 0.0039, and 0.0020 vs baseline), respectively (**Figure 4**).^lxi^ All 15 patients in the 400-mg BID group, all 9 patients in the 600-mg BID group, and 9 of 10 patients in the 800-mg QD group achieved hemolysis control.^lxii^

**Figure 3.**
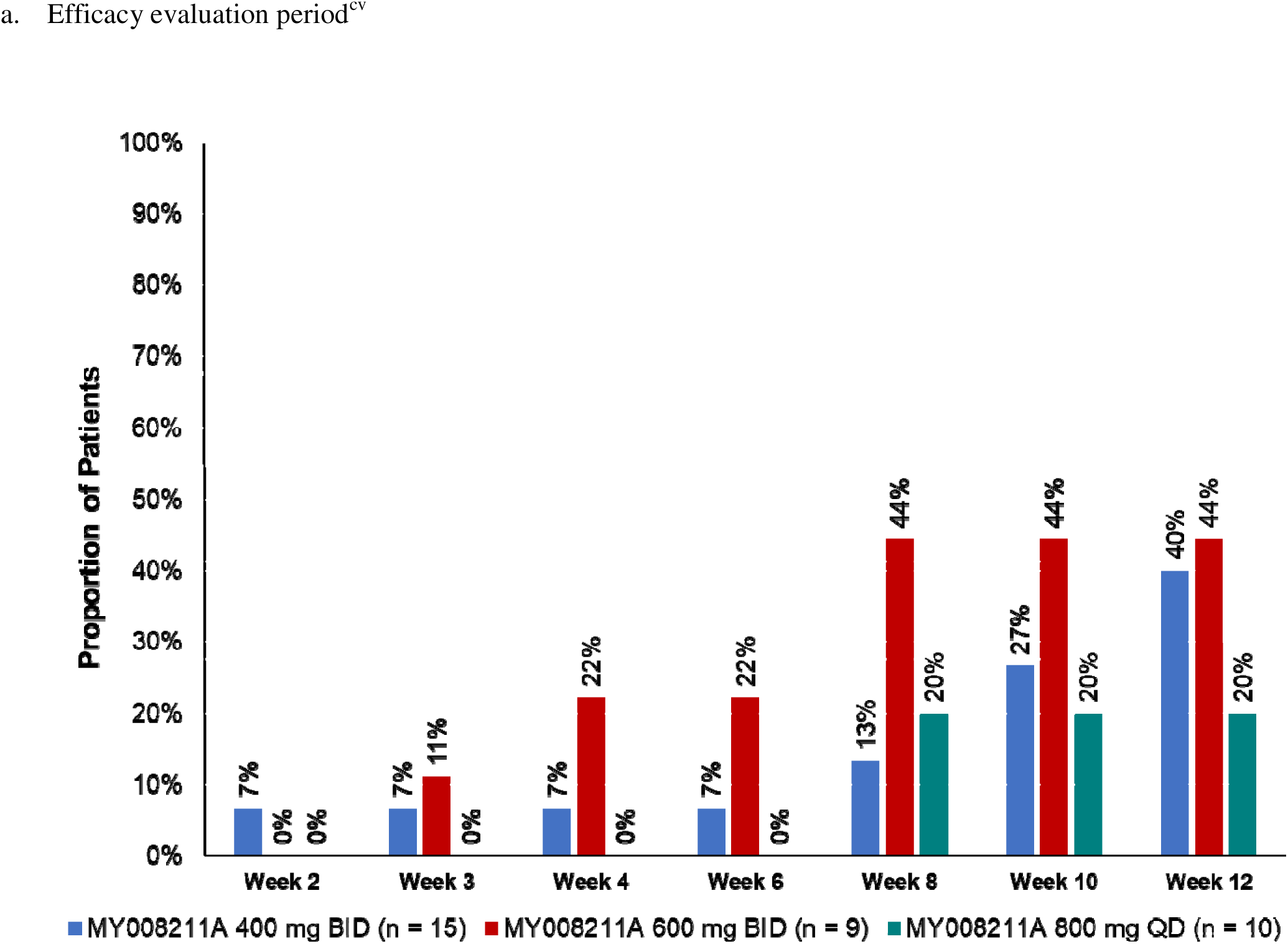
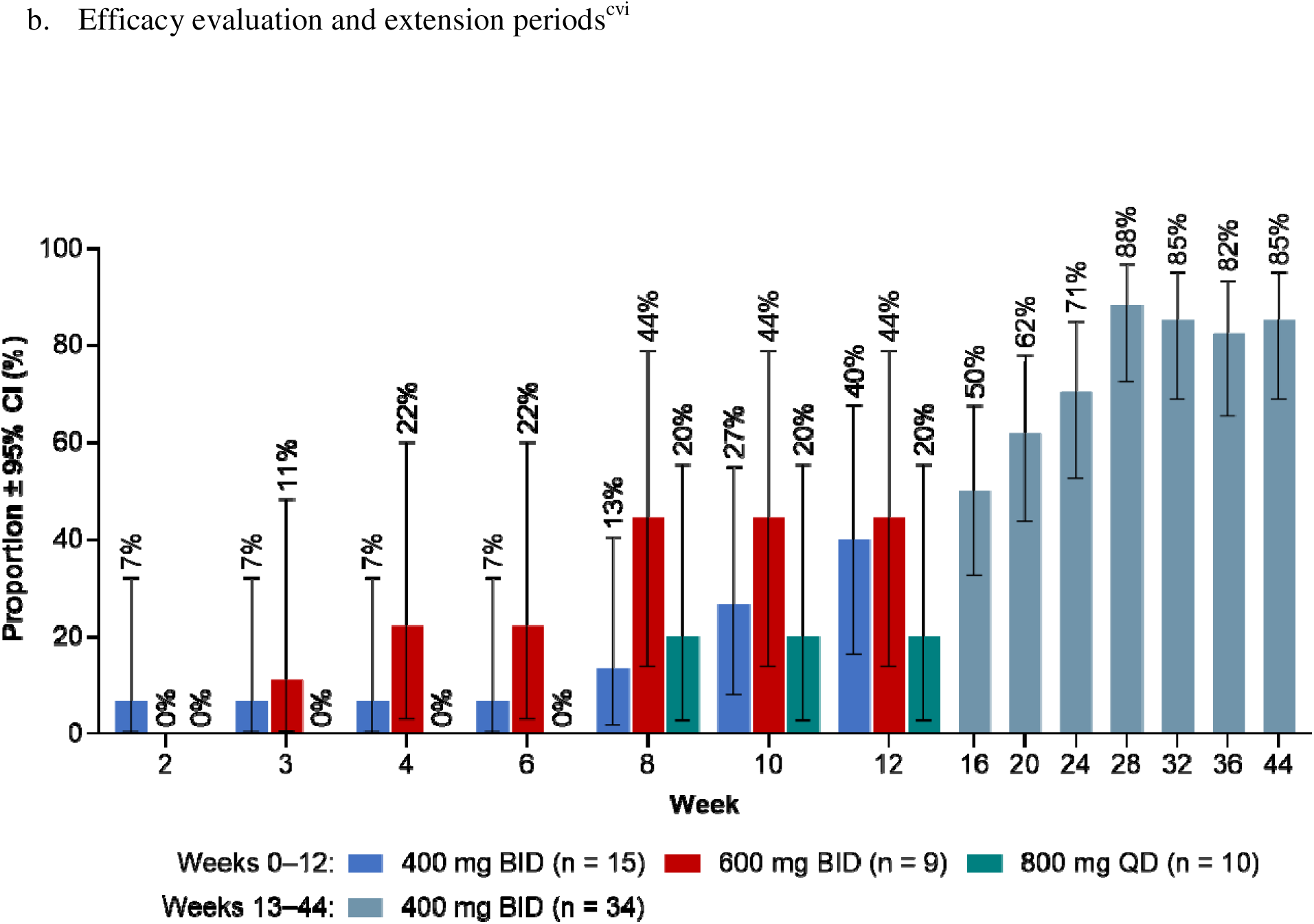
Proportion of patients without RBC transfusion with hemoglobin concentration ≥120 g/L. BID, twice daily; CI, confidence interval; QD, once daily; RBC, red blood cell.

**Figure 4.**
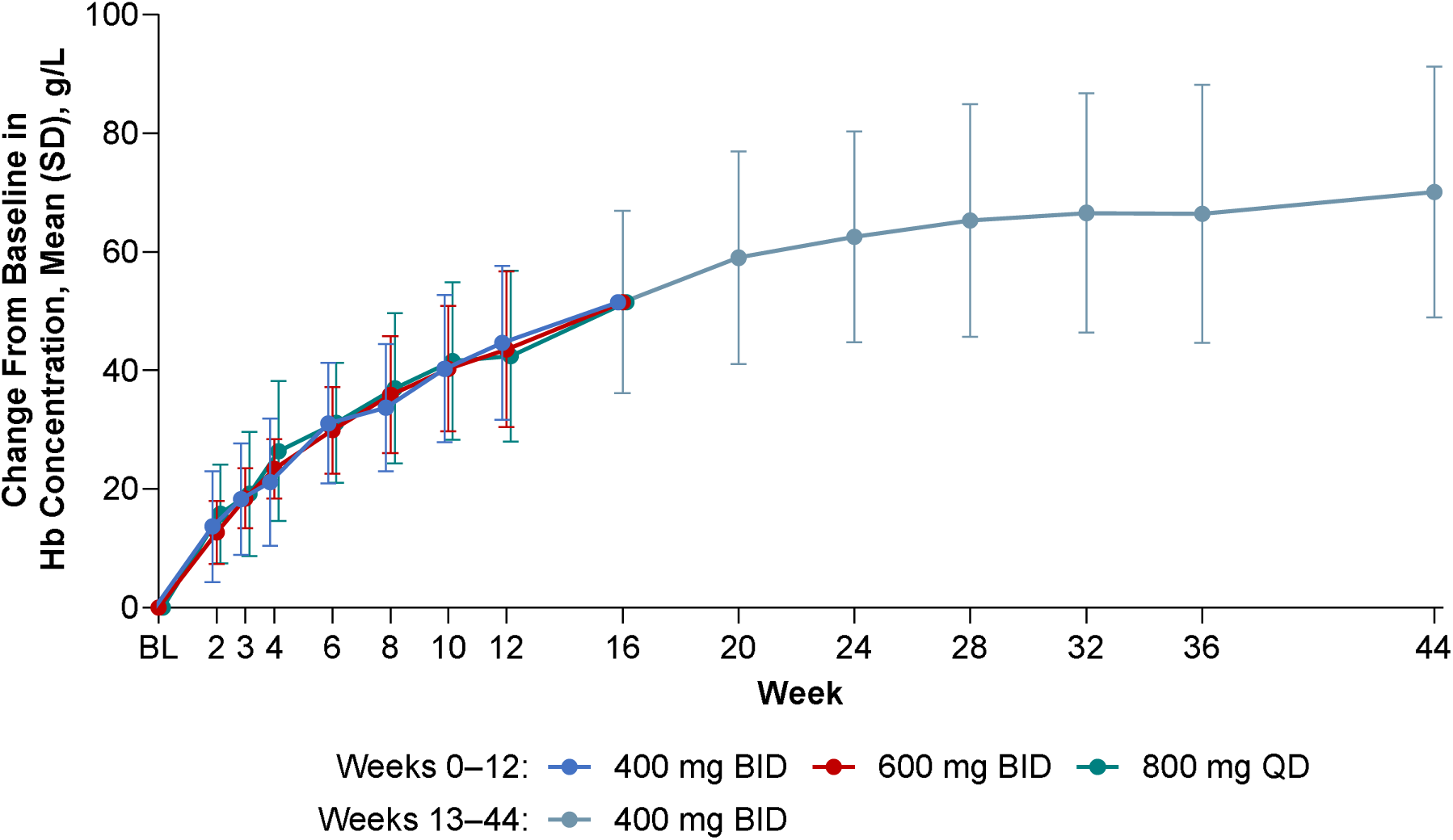
Changes from baseline in hemoglobin concentration^cvii^. BID, twice daily; Hb, hemoglobin; QD, once daily; SD, standard deviation.

The changes from baseline in reticulocyte counts and indirect bilirubin levels were significant in all groups.^lxiii^ Mean (SD) changes in reticulocyte counts were −118.27 (67.043), −84.26 (67.404), and −90.89 (67.921) × 10^9^/L in the 400-mg BID, 600-mg BID, and 800-mg QD groups (*P* =0.0001, 0.0039, and 0.0059 vs baseline), respectively.^lxiv^ Mean (SD) changes in indirect bilirubin levels were −15.032 (9.5471), −14.511 (7.0085), and −11.796 (14.1008) µmol/L in the 400-mg BID, 600-mg BID, and 800-mg QD groups (*P* <0.0001, 0.0039, and 0.0039 vs baseline), respectively.^lxv^ Fatigue, as measured by the FACIT-F score, improved from baseline by a mean (SD) of 9.7 (11.8) points (higher scores indicate reduced fatigue)^lxvi^ with 400 mg BID, 3.2 (6.1) points with 600 mg BID, and 8.7 (7.6) points with 800 mg QD (**Table 2**).^lxvii^ Mean (SD) changes from baseline in PNH RBC clone levels were 34.22% (15.304) with 400 mg BID, 29.58% (18.298) with 600 mg BID, and 35.71 (16.099) with 800 mg QD.^lxviii^

**Table 2.** Changes from baseline to weeks 12 and 44 in secondary endpoints^cxiii^.

|  | 0–12 weeks |  |  | 13–44 weeks<sup>cxiv</sup> |
| --- | --- | --- | --- | --- |
|  | 400 mg BID<br>n□=□15 | 600 mg BID<br>n□=□9 | 800 mg QD<br>n□=□10 | 400 mg BID<br>n□= 34 |
| **Hemoglobin, g/L** |  |  |  |  |
| Value at week 12 or 44 | 115.3 (14.29) | 119.6 (21.41) | 107.7 (14.38) | 140.6 (16.26) |
| Change | 44.7 (12.98) | 43.6 (13.21) | 42.4 (14.40) | 70.1 (21.17) |
| *P*-value | <0.0001 | 0.0039 | 0.0020 | <0.0001 |
| **LDH, U/L** |  |  |  |  |
| Value at week 12 or 44 | 246.487 (73.0242) | 236.556 (54.7908) | 320.164 (179.1376) | 258.844 (62.6572) |
| Change | –1672.27 (811.531) | –1527.44 (418.785) | –1332.97 (392.153) | –1540.82 (621.150) |
| *P*-value | <0.0001 | 0.0039 | 0.0020 | <0.0001 |
| **Reticulocyte count, 10<sup>9</sup>/L** |  |  |  |  |
| Value at week 12 or 44 | 116.44 (68.534) | 109.89 (25.196) | 109.12 (42.117) | 118.86 (40.215) |
| Change | –118.27 (67.043) | –84.26 (67.404) | –90.89 (67.921) | –94.90 (72.252) |
| *P*-value | 0.0001 | 0.0039 | 0.0059 | <0.0001 |
| **Indirect bilirubin, μmol/L** |  |  |  |  |
| Value at week 12 or 44 | 13.288 (7.6336) | 8.822 (2.3921) | 12.801 (4.0064) | 15.148 (6.5894) |
| Change | −15 (9.5) | −15 (7.0) | −12 (14.1) | −11 (10.5) |
| *P*-value | <0.0001 | 0.0039 | 0.0039 | <0.0001 |
| **RBC clone levels, %\*** |  |  |  |  |
| Week 12 value<sup>cxv</sup> | 86.454 (8.0867) | 85.857 (7.9940) | 84.557 (12.0589) | — |
| Change | 34.219 (15.3038) | 29.580 (18.2979) | 35.707 (16.0991) | — |
| *P*-value | <0.0001 | 0.0078 | 0.0020 | — |
| **FACIT-F score\*<sup>cxvi</sup>** |  |  |  |  |
| Week 12 value | 43.5 (5.26) | 42.8 (5.26) | 45.1 (4.63) | — |
| Change | 9.7 (11.8) | 3.2 (6.1) | 8.7 (7.6) | — |
| *P*-value | 0.0054 | 0.2031 | 0.0059 | — |

\*Collected through week 12.
Changes from baseline are expressed as mean (SD). *P*-values are for the comparison of week 12 or 44 vs baseline. As efficacy over the extension period was evaluated post hoc, *P*-values should be considered nominal.
BID, twice daily; FACIT-F, Functional Assessment of Chronic Illness Therapy–Fatigue; LDH, lactate dehydrogenase; QD, once daily; RBC, red blood cell; SD, standard deviation.

Efficacy over the 32-week extension period was analyzed post hoc. At week 44, all 34 patients treated with MY008211A maintained the primary endpoint result of an increase in hemoglobin concentration of ≥20 g/L from baseline (**Figure 2b**).^lxix^ Additionally, at week 44, 29 of 34 patients (11 of 15 patients originally in the 400-mg BID group, 8 of 9 patients originally in the 600-mg BID group, and all 10 patients originally in the 800-mg QD group) had a hemoglobin concentration ≥120 g/L (**Figure 3b**).^lxx^

**Table 2** shows changes from baseline to week 44 in hemoglobin, LDH, reticulocyte count, and indirect bilirubin; all treatment groups maintained differences vs baseline over the extension.^lxxi^ Thirty-two patients (13 originally in the 400-mg BID group, 9 originally in the 600-mg BID group, and 10 originally in the 800-mg QD group) maintained hemolysis control at week 44.^lxxii^ All patients completed 44 weeks of treatment without RBC transfusions.^lxxiii^

### Safety

Over the 12-week efficacy evaluation period, 93%, 89%, and 100% of patients in the 400-mg BID, 600-mg BID, and 800-mg QD groups, respectively, experienced adverse events (AEs); over the extension period, 88% of patients experienced an AE.^lxxiv^ Overall, AEs were mild to moderate in intensity. Five patients experienced 8 grade 3 AEs over the efficacy evaluation period, and 8 patients experienced 10 grade 3 events over the extension. Three patients experienced 3 grade 4 AEs over the efficacy evaluation period, while no grade 4 events occurred during the extension. Two (13%) patients in the 400-mg BID group had 2 serious AEs (SAEs), 2 (22%) patients in the 600-mg BID group had 2 SAEs, and 3 (30%) patients in the 800-mg QD group had 6 SAEs.^lxxv^ These SAEs included respiratory tract infection (2 patients in the 400-mg BID group and 1 patient in the 800-mg QD group), both infection and breakthrough hemolysis in 1 patient in the 800-mg QD group, both respiratory tract infection and breakthrough hemolysis in 1 patient in the 800-mg QD group, radius fracture in 1 patient in the 600-mg BID group, breakthrough hemolysis in 1 patient in the 600-mg BID group, and ruptured cerebral aneurysm in 1 patient in the 800-mg QD group.^lxxvi^ During the entirety of the study (including efficacy observation and the extension), the most frequent AEs across all dosage groups were headache (n[=[15 of 34 patients) and upper respiratory infection (n[=[11; **Table 3**).^lxxvii^ Nine of 15, 6 of 9, and 6 of 10 patients in the 400-mg BID, 600-mg BID, and 800-mg QD groups experienced an infection; of the total 29 occurrences of an infection event, 18 were either respiratory tract infection or upper respiratory infection.^lxxviii^

**Table 3.** Safety results^cxvii^.

|  | **400 mg BID**<br>**n□=□15**<br>**Weeks 0–12** |  | **600 mg BID**<br>**n□=□9**<br>**Weeks 0–12** |  | **800 mg QD**<br>**n□=□10**<br>**Weeks 0–12** |  | **Total**<br>**N□=□34**<br>**Weeks 0–12** |  | **400 mg BID**<br>**n□=□34**<br>**Weeks 13–44<sup>cxviii</sup>** |  |
| --- | --- | --- | --- | --- | --- | --- | --- | --- | --- | --- |
|  | **e/n** | **% of N** | **e/n** | **% of N** | **e/n** | **% of N** | **e/n** | **% of N** | **e/n** | **% of N** |
| **AE** | 42/14 | 93.3 | 31/8 | 88.9 | 54/10 | 100.0 | 127/32 | 94.1 | 127/30 | 88.2 |
| **AE judged as related to the study drug** | 19/11 | 73.3 | 20/7 | 77.8 | 20/7 | 70.0 | 59/25 | 73.5 | 31/16 | 47.1 |
| **SAE** | 0 | 0 | 0 | 0 | 4/2 | 20.0 | 4/2 | 5.9 | 6/5 | 14.7 |
| **SAER** | 0 | 0 | 0 | 0 | 0 | 0 | 0 | 0 | 1/1 | 2.9 |
| **Grade 3 AE** | 0 | 0 | 4/3 | 33.3 | 4/2 | 20.0 | 8/5 | 14.7 | 10/8 | 23.5 |
| **Grade 4 AE** | 1/1 | 6.7 | 1/1 | 11.1 | 1/1 | 10.0 | 3/3 | 8.8 | 0 | 0 |
| **Grade 5 AE** | 0 | 0 | 0 | 0 | 0 | 0 | 0 | 0 | 0 | 0 |
| **Headache** | 8/8 | 53.3 | 3/2 | 22.2 | 5/5 | 50.0 | 16/15 | 44.1 | 0 | 0 |
| **URTI** | 3/3 | 20.0 | 1/1 | 11.1 | 2/2 | 20.0 | 6/6 | 17.6 | 8/6 | 17.6 |
| **ALT increased** | 1/1 | 6.7 | 2/2 | 22.2 | 3/3 | 30.0 | 6/6 | 17.6 | 6/5 | 14.7 |
| **Arthralgia** | 4/4 | 26.7 | 0 | 0 | 1/1 | 10.0 | 5/5 | 14.7 | 5/5 | 14.7 |
| **WBC decreased** | 2/1 | 6.7 | 5/4 | 44.4 | 7/3 | 30.0 | 14/8 | 23.5 | 6/4 | 11.8 |
| **AST increased** | 1/1 | 6.7 | 0 | 0 | 4/4 | 40.0 | 5/5 | 14.7 | 5/5 | 14.7 |
| **Influenza-like illness** | 2/2 | 13.3 | 0 | 0 | 2/2 | 20.0 | 4/4 | 11.8 | 4/4 | 11.8 |
| **Hyperlipidemia** | 2/2 | 13.3 | 0 | 0 | 2/2 | 20.0 | 4/4 | 11.8 | 3/3 | 8.8 |
| **Hypokalemia** | 1/1 | 6.7 | 2/2 | 22.2 | 2/2 | 20.0 | 5/5 | 14.7 | 8/4 | 11.8 |

e/n indicates events/number of patients with event.
AE, adverse event; ALT, alanine aminotransferase; AST, aspartate aminotransferase; BID, twice daily; QD, once daily; SAE, serious adverse event; SAER, severe adverse event related to study drug; URTI, upper respiratory tract infection; WBC, white blood cells.

Over the 44 weeks of treatment, AEs judged as related to the study drug were experienced by 12 (80%), 8 (89%), and 8 (80%) patients originally randomized to the 400-mg BID, 600-mg BID, and 800-mg QD groups, respectively, including 24, 30, and 36 events.^lxxix^ Of these, 31 events in 16 patients occurred during the extension period. **^lxxx^** Only 1 SAE was judged as possibly related to the study drug: a grade 3 serious respiratory tract infection in the 800-mg QD group that occurred during the extension period.^lxxxi^ There were no deaths^lxxxii^ and no treatment discontinuations.^lxxxiii^ In total, there were 4 episodes of breakthrough hemolysis. There was 1 instance of breakthrough hemolysis (7%) in the 400-mg BID group, 1 in the 600-mg BID group (11%), and 2 in the 800-mg QD group (20%).^lxxxiv^ The patient in the 400-mg BID group developed breakthrough hemolysis on study day 133 and experienced respiratory tract infection before this event. The patient in the 600-mg BID group developed breakthrough hemolysis after receiving the study drug for 247 days and experienced fatigue, diarrhea, and lung infection before the event. Two breakthrough hemolysis events that developed in patients in the 800-mg QD group were reported as SAEs, including in 1 patient who developed breakthrough hemolysis accompanied with infection 74 days after taking the study drug and another patient who developed breakthrough hemolysis 106 days after taking the study drug, accompanied by dose omission and fatigue. All the breakthrough hemolysis events were considered unlikely to be related to the study drug, and all events resolved There were no major adverse cardiac events or thrombotic episodes, and there were no meningococcal infections.

## Discussion

In this multicenter, open-label study of 3 dosages of MY008211A tablets, results over the 12-week efficacy evaluation period among 34 patients with PNH and signs of active hemolysis showed that all patients achieved increases in hemoglobin concentration of ≥20 g/L from baseline by the end of treatment without the need for RBC transfusions. All patients maintained these improvements for the duration of the 32-week extension. Improvements in secondary endpoints, including fatigue, were of similar magnitude across dosage groups. All but 1 patient achieved hemolysis control.^lxxxv^

Complement inhibitors represent the usual standard-of-care treatment for PNH.^11lxxxvi^ In the pivotal phase 3 trial of eculizumab (N = 87), treatment with eculizumab rapidly reduced hemolysis, as indicated by decreased LDH.^lxxxvii^ After 26 weeks, 49% of patients receiving eculizumab had stabilized hemoglobin levels vs 0% in the placebo group (*P* <0.001), and the median of packed red cells was 0 and 10 units per patient, respectively (*P* <0.001).^12lxxxviii^ Additional complement inhibitors have been evaluated and added to the therapeutic armamentarium after demonstrating successful amelioration of hemolysis. These include additional C5 inhibitors, such as ravulizumab, which was found to be noninferior to eculizumab in inducing LDH normalization (54% vs 49%) and preventing the need for transfusion (74% vs 66%) in a 26-week trial that included 246 patients.^13lxxxix^ The C3 inhibitor pegcetacoplan was found to be superior to eculizumab for change in hemoglobin concentration from baseline to week 16 (mean difference, 3.84 g/dL; *P* <0.001).^14xc^ The complement factor D inhibitor danicopan improved hemoglobin levels vs placebo in a trial of 86 patients over 12 weeks; patients who switched from placebo to danicopan then experienced improved hemoglobin levels.^15xci^ Iptacopan, a complement factor B inhibitor, was associated with a greater proportion of patients having an increase in hemoglobin concentration of at least 2 g/dL compared with anti-C5 therapy in one phase 3 trial (N = 97) over 24 weeks; in another phase 3 trial, 92% of complement inhibitor–naive patients had a hemoglobin increase of at least 2 g/dL without RBC transfusion (N = 40).^16xcii^

Nevertheless, these effective agents have drawbacks, including intravenous dosing, and risk of infection with eculizumab and other agents^9xciii^, and risk of iatrogenic extravascular hemolysis and persistent anemia with C5 inhibition.^5xciv^ Complement inhibitors have been found to increase the risk of viral infections, and a large pharmacovigilance study using the United States Food & Drug Administration Adverse Event Reporting System found that this increase was particularly notable with C3 inhibitors.^7xcv^ In the study reported here, the orally administered complement factor B inhibitor MY008211A yielded significant increases in hemoglobin and RBC and decreases in LDH, while observed AEs were predominantly mild to moderate in severity, and infections generally consisted of respiratory infections. There were no discontinuations over the treatment period and no thrombotic events.^xcvi^ The improved hemolytic outcomes in this study were also accompanied by clinical benefits, as indicated by a clinically important 9.7-point increase in FACIT-F score in the 400-mg BID group.^17xcvii^

Limitations of this research include that this patient population was almost entirely Han Chinese; clinical presentation of PNH can differ across ethnic groups, so these findings may not be generalizable to a broader multiethnic population.^1xcviii^ The lack of a randomized control group may be considered a limitation, although the study design was intended to provide for appropriate comparisons with historical controls. The size of this phase 2 trial is also a limitation, and larger studies will need to be undertaken to further characterize the safety and efficacy of MY008211A tablets.

Taken together, the safety and efficacy data from this phase 2 study including a 12-week treatment period and 32-week extension suggest that MY008211A tablets may benefit patients with PNH. Further evaluation is warranted, including determination of whether the 400-mg BID dosage is best suited for clinical development.

## Data Availability

All data produced in the present study are available upon reasonable request to the authors

## Acknowledgments

This study was funded by Wuhan Createrna Science and Technology Co., Ltd. Editorial and writing support were provided by Rob Coover, MPH, of Red Nucleus, and funded by Createrna.

## Authorship contributions

F.Z., C.X., R.K., and Y.C. designed the research. L.Y., M.C., H.T., L.Z., H.C., X.L., Z.S., G.X., and N.G. performed the research, including patient recruitment, treatment, and follow-up. F.Z., Y.C., and R.K. provided study oversight and scientific governance. X.L. was responsible for medical monitoring and safety assessment. R.X., C.T., L.L., X.G., Y.Z., and W.C.W. managed project coordination and study operations. L.Y., M.C., H.T., C.X., and F.Z. analyzed and interpreted the data. L.Y., M.C., and H.T. wrote the manuscript. F.Z., C.X., R.K., G.R., C.D.C., B.H., and Y.C. critically revised the manuscript for important intellectual content. All authors reviewed and approved the final manuscript.^xcix^

---

**Supplemental Figure 1.**
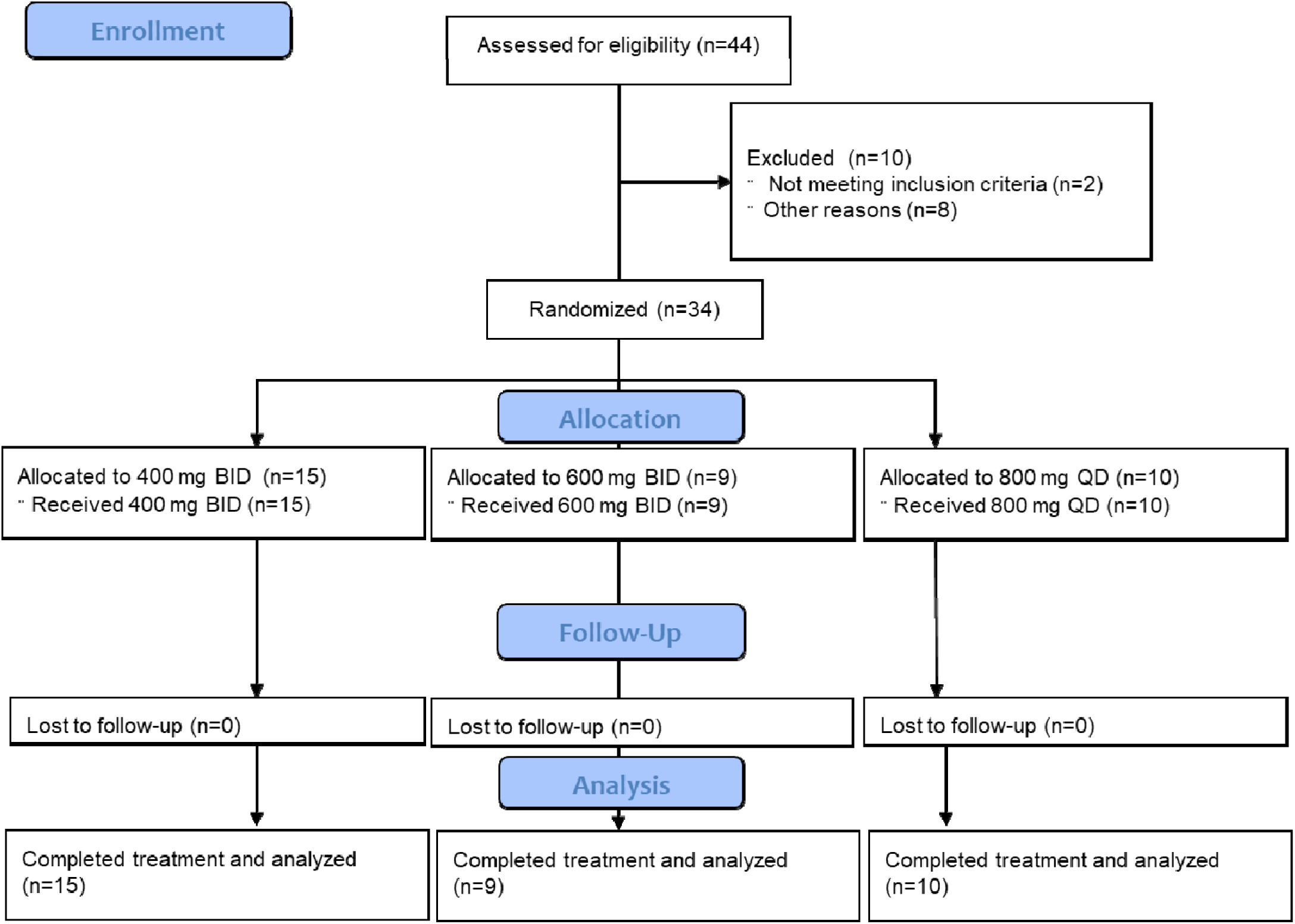
Patient disposition^cxix^.

## Footnotes

i 2024ASH_Abstract_draft.07.11.2024

ii 2024ASH_Abstract_draft.07.11.2024

iii Lian_responses_260130 p1

iv Translated_MY008211A-PNH-2-01_Post-hoc_20250429_20250530 p4

v Munoz-Linare EurJHaematol 2014 p1 col1 para1-2

vi Oliver JBloodMed 2023 recommend p4 para5; Panse TransfusMedHemother 2024 p3 col2 para3; Bodo AdvTher 2023 CP5 p14 col1 para1

vii translated_MY008211A-PNH-2-01_TLFs_20250415-tables14.3.1.1-tables14.3.2.10 AE 20250530 p3-8; Translated_MY008211A-PNH-2-01_Post-hoc 20250429 20250530 p17, p27, p41-2, p63-4

viii https://www.hopkinsmedicine.org/kimmel-cancer-center/cancers-we-treat/blood-bone-marrow-cancers/paroxysmal-nocturnal-hemoglobinuria-pnh

ix https://www.createrna.com/en/news-activity/news-center/news/46

x MY008-PNH-Phase2 protocol synopsis-V2.1-draft translation, page 7

xi MY008-PNH-Phase2 protocol synopsis-V2.1-draft translation, page 11. Country where trial was conducted was communicated to us verbally by client; MY008-PNH-Phase2 protocol synopsis-V2.1-draft translation, pages 12 & 14

xii MY008-PNH-Phase2 protocol synopsis-V2.1-draft translation, pages 12 & 14; MY008211A-PNH-Phase2-CSR-Addendum-2025.06.07, p8, para 1

xiii MY008-PNH-Phase2 protocol synopsis-V2.1-draft translation, page 18

xiv MY008-PNH-Phase2 protocol synopsis-V2.1-draft translation, pages 17-18

xv 2024_ASH_Abstract_SafetyTLFs.07.11.2024, page 1. We don’t have directly supportive materials, eg, a disposition chart/table, but all the efficacy and safety tables give 34 total, broken down as 15 with 400, 9 with 600, and 10 with 800, and there were no DCs.

xvi Translated_MY008211A-PNH-Phase2-CSR-Addendum-2025.06.07 p9

xvii 2024_ASH_Abstract_EfficacyTLFs.07.11.2024, page 1

xviii Translated_MY008211A-PNH-2-91_Post-hoc_20250429 20250530 p3

xix Translated_MY008211A-PNH-Phase2-CSR-Addendum-2025.06.07 p17

xx Translated_MY008211A-PNH-Phase2-CSR-Addendum-2025.06.07 p12

xxi Hill NatRevDisPrimers 2021 PNH p7 para2, 3, 4

xxii Munoz-Linare EurJHaematol 2014 p1 col1 para1-2

xxiii Richards EurJHematol 2021 abs p3

xxiv Panse TransfusMedHemother 2024 p6 col2 para3

xxv Panse TransfusMedHemother 2024 p7 Fig1

xxvi Oliver JBloodMed 2023 recommend p4 para5; Panse TransfusMedHemother 2024 p3 col2 para3; Bodo AdvTher 2023 CP5 p14 col1 para1

xxvii Oliver JBloodMed 2023 p5-6 Table 1

xxviii Zhong FrontPharmacol 2025 infection p6 col2 para1

xxix Bodo AdvTher 2023 CP5 p1 key summary points

xxx Oliver JBloodMed 2023 p2 para2

xxxi Markiewicz BBMT 2020 PNH transplant p3 col1 para2, col2 para1

xxxii Cancado HematolTransfusCellTher 2020 PNH consensus p3 col2 HCT section

xxxiii Lian_responses_260130

xxxiv https://clinicaltrials.gov/study/NCT06050226

xxxv MY008-PNH-Phase2 protocol synopsis-V2.1-draft p6; number of centers added by client

xxxvi MY008-PNH-Phase2 protocol synopsis-V2.1-draft p14-15 Inclusion and Exclusion criteria

xxxvii Lu Intractable Rare Dis Res. 2022 p2 col1 para1

xxxviii MY008-PNH-Phase2 protocol synopsis-V2.1-draft translation p14

xxxix MY008-PNH-Phase2 protocol synopsis-V2.1-draft p 28-29 #1, #7

xl Translated_MY008211A-PNH-Phase2-CSR-Addendum-2025.06.07 p7-8

xli Translated_MY008211A-PNH-Phase2-CSR-Addendum-2025.06.07 p7-8

xlii MY008-PNH-Phase2 protocol synopsis-V2.1-draft translation p18

xliii MY008-PNH-Phase2 protocol synopsis-V2.1-draft translation p18-19

xliv Translated_MY008211A-PNH-2-01_Post-hoc_20250429 20250530 p3, 4, 6, 7, 13-17, 23-28, 30-31, 37-42, 48-53, 59-64, 65-66

xlv Translated_MY008211A-PNH-2-01_Post-hoc_20250429 20250530 p30, MY008-PNH-Phase2 protocol synopsis-V2.1-draft translation p18

xlvi Translated_MY008211A-PNH-2-01_Post-hoc_20250429 20250530 p65

xlvii mY008-PNH-Phase2 protocol synopsis-V2.1-draft translation p22

xlviii The client added the “45%.” I haven’t found it in source documents

xlix mY008-PNH-Phase2 protocol synopsis-V2.1-draft translation p23

l mY008-PNH-Phase2 protocol synopsis-V2.1-draft translation p22-23

li Lian-responses-251209 in Correspondence

lii 2024_ASH_Abstract_demographicsTLFs.077.22.2024 p1

liii 2024_ASH_Abstract_demographicsTLFs.077.22.2024 p1-2

liv Translated_MY008211A-PNH-2-01_TLFs_20250415-tables14.1.1.1-tables14.1.3.8 Baseline 20250530 p5; Lian-responses-251209

lv Translated_MY008211A-PNH-2-01_TLFs_20250415-tables14.1.1.1-tables14.1.3.8 Baseline 20250530 p7

lvi Translated_MY008211A-PNH-Phase2-CSR-Addendum-2025.06.07 p9

lvii Translated_MY008211-PNH-2-01_TLFs_20250415-tables14.3.1.1-tables14.3.2.10 AE 20250530 p1

lviii 2024_ASH_Abstract_EfficacyTLFs.07.11.2024 p1

lix 2024_ASH_Abstract_EfficacyTLFs.07.11.2024 p1

lx Translated_MY008211A-PNH-2-01_Post-hoc 20250429 20250530 p6

lxi Translated_MY008211A-PNH-2-01_Post-hoc 20250429 20250530 p12, MY008-PNH-Phase2 protocol synopsis-V2.1-draft translation p6 for units

lxii 2024_ASH_Abstract_EfficacyTLFs.07.11.2024 p12

lxiii 2024_ASH_Abstract_EfficacyTLFs.07.11.2024 p14,16

lxiv Translated_MY008211A-PNH-2-01_Post-hoc 20250429 20250530 p37; MY008-PNH-Phase2 protocol synopsis-V2.1-draft translation p14 for units

lxv Translated_MY008211A-PNH-2-01_Post-hoc 20250429 20250530 p59; translated_MY008211A-PNH-2-01_Post-hoc figures_20250530 p13 for units

lxvi pg 31, #21 of MY008-PNH-Phase2 protocol synopsis-V2.1-draft translation

lxvii 2024_ASH_Abstract_EfficacyTLFs.07.11.2024 p18

lxviii table2update p3

lxix Translated_MY008211A-PNH-2-01_Post-hoc 20250429 20250530 p4

lxx Translated_MY008211A-PNH-2-01_Post-hoc 20250429 20250530 p7

lxxi Translated_MY008211A-PNH-2-01_Post-hoc 20250429 20250530 p17, p27, p41-2, p63-4

lxxii Translated_MY008211A-PNH-2-01_Post-hoc 20250429 20250530 p31

lxxiii Translated_MY008211A-PNH-2-01_Post-hoc 20250429 20250530 p66

lxxiv Table3 p1

lxxv Translated_MY008211A-PNH-Phase2-CSR-Addendum-2025.06.07 p12 Table 6

lxxvi Translated_MY008211A-PNH-Phase2-CSR-Addendum-2025.06.07 19-28

lxxvii Translated_MY008211A-PNH-Phase2-CSR-Addendum-2025.06.07 p14 Table 6-2

lxxviii Translated_MY008211A-PNH-2-01_TLFs_20250415-tables14.3.1.1-tables14.3.2.10 AE 20250530 p4

lxxix Translated_MY008211A-PNH-2-01_TLFs_20250415-tables14.3.1.1-tables14.3.2.10 AE 20250530 p2

lxxx Table3 from MY008211a-pnh-2-01 ad hoc

lxxxi Translated_MY008211A-PNH-Phase2-CSR-Addendum-2025.06.07 p22 bullet, p24 para3; Table3

lxxxii Translated_MY008211A-PNH-Phase2-CSR-Addendum-2025.06.07 p18 6.3.1.1.

lxxxiii Translated_MY008211A-PNH-Phase2-CSR-Addendum-2025.06.07 p11 6.2.1.

lxxxiv Translated_MY008211A-PNH-Phase2-CSR-Addendum-2025.06.07 p15

lxxxv 2024_ASH_Abstract_EfficacyTLFs.07.11.2024 p12

lxxxvi Brodsky Blood 2021 HowITreatPNH p1 col1 para3

lxxxvii Hillmen NEJM 2006 Eculizumab p4 col2 last para

lxxxviii Hillmen NEJM 2006 Eculizumab p2 col1 para4, p3 col1 para3, p4 Table 1, p6 Table 2

lxxxix Lee Blood 2019 Ravulizumab p5 col1 para2, para3

xc Hillmen NEnglJMed 2021 Pegcetacoplan p4 col1 para3

xci Kulasekararaj Blood 2025 danicopan p3 Table 1, p4 Table 2

xcii De Latour_NEJM_2024_994_iptacopan p3 ol2 para4, p4 col1 para3-4

xciii Cancado HematolTransfusCellTher 2021 guidelines p5 col1 para 1,2

xciv Oliver JBloodMed 2023 recommend p4 para5

xcv Zhong FrontPharmacol 2025 infection p6 col2 para1

xcvi translated_MY008211A-PNH-2-01_TLFs_20250415-tables14.3.1.1-tables14.3.2.10 AE 20250530 p3-8

xcvii Cella JPtReportedOutcomes 2023 fatigue p5 col1 para1

xcviii Hill NatRevDisPrimers 2021 PNH p3 para2

xcix Lian_responses_260130 p2

c Lian_responses_260130 p2

ci MY008-PNH-Phase2 protocol synopsis-V2.1-draft translation p12-13

cii MY008-PNH-Phase2 protocol synopsis-V2.1-draft translation p13

ciii translated_MY008211A-PNH-2-01_Post-hoc_20250429 20250530

civ figure_2b en-US

cv pgs 5-6 of translated_MY008211A-PNH-2-01_Post-hoc_20250429 20250530

cvi figure_3b en-US

cvii figure_4 en-US

cviii 2024_ASH_Abstract_demographicsTLFs.07.22.2024

cix Translated_MY008211A-PNH-2-01_TFLs_20250415-tables14.1.1.1-tables14.1.3.8 Baseline 20250530 p13

cx Translated_MY008211A-PNH-2-01_TFLs_20250415-tables14.1.1.1-tables14.1.3.8 Baseline 20250530 p18

cxi Translated_MY008211A-PNH-2-01_TFLs_20250415-tables14.1.1.1-tables14.1.3.8 Baseline 20250530 p17

cxii Translated_MY008211A-PNH-2-01_TFLs_20250415-tables14.1.1.1-tables14.1.3.8 Baseline 20250530 p19

cxiii 2024_ASH_Abstract_EfficacyTLFs07.11.2024 p4, 9, 14, 16

cxiv Table2 from MY008211a-pnh-2-01 ad hoc

cxv Table2update p3

cxvi 2024_ASH_Abstract_EfficacyTLFs.07.11.2024, p18 and Table2update p3

cxvii Translated_MY008211A-PNH-Phase2-CSR-Addendum-2025.06.07 p12 Table 6-1, p14 Table 6-2

cxviii Table3 from MY008211a-pnh-2-01 ad hoc

cxix TranslatedTranslate_MY008211A-PNH-Phase2-CSR-Addendum-2025.06.07 p9p33 Table 5-1

